# Decreases and Pronounced Geographic Variability in Antibiotic Prescribing in Medicaid

**DOI:** 10.1101/2023.02.04.23285480

**Authors:** Alexia G. Aguilar, Priscilla C. Canals, Kimberly A. Miller, Brian J. Piper

## Abstract

**Background:** Antibiotic resistance is a persistent and growing concern. Our objective was to analyze antibiotic prescribing data in the United States (US) to identify trends in the Medical Expenditure Panel System (MEPS) and any state-level disparities to Medicaid patients.

**Methods:** We obtained total MEPS prescriptions for eight antibiotics from 2013 to 2020. We extracted prescribing rates per 1,000 Medicaid enrollees for two years, 2018 and 2019, for four broad spectrum (azithromycin, ciprofloxacin, levofloxacin, and moxifloxacin) and four narrow spectrum (amoxicillin, cephalexin, doxycycline, and trimethoprim/sulfamethoxazole) antibiotics.

**Results:** Antibiotics in MEPS decreased from 2013 to 2020 by −38.7% with a larger decline for broad (−53.7%) than narrow spectrum (−23.5%). Antibiotic prescriptions decreased from 2018 to 2019 and by −6.7% when correcting for the number of Medicaid enrollees. Amoxicillin was the predominant antibiotic followed by azithromycin, cephalexin, trimethoprim/sulfamethoxazole, doxycycline, ciprofloxacin, levofloxacin, and moxifloxacin. Substantial geographic variation in antibiotic prescribing existed with 2.8-fold between the highest (Kentucky = 855/1,000) and lowest (Oregon = 299) states. The South prescribed 52.2% more antibiotics (580/1,000) than the West (381/1,000). There were significant correlations across states in antibiotic prescribing.

**Conclusions & Relevance:** This study identified sizable disparities by geography in prescribing rates of eight antibiotics with over three-fold state level differences. The South had the highest prescribing rates among all the regions. Areas with high antibiotic prescribing rates, particularly for outpatients, may benefit from programs to reduce potentially unnecessary prescribing. Further analysis of state level Medicaid or prescribing policies is needed to identify reasons for the variation in prescribing rates.

---

Antibiotic resistance is become one of the most pressing threats to public health today. According to the World Health Organization, increasing antibiotic resistance is one of the biggest threats to global health, food security, and the development of human defenses today.^1^ Overuse and inappropriate prescribing of antibiotics has been identified in a variety of healthcare settings.^2-5^ According to a 2019 report from the Centers for Disease Control and Prevention, more than 2.8 million antibiotic-resistant infections occur in the United States (US) each year, and more than 35,000 people died as a result.^6^ Most of these antibiotic prescriptions were for respiratory infections, commonly caused by viruses, which do not respond to antibiotics.^7^ Antibiotics are often prescribed unnecessarily,^8,9^ and 30-50% of prescriptions for them are not associated with an indication.^10-12^ Sulfonamides and urinary anti-infective agents are the classes most likely to be prescribed without documentation.^11^ Up to 25% of antibiotics prescribed in the outpatient setting to Medicaid beneficiaries were not associated with a provider visit and therefore were not screened by existing antimicrobial stewardship systems.^13^ Further, among 298 million prescriptions filled by 53 million Medicaid patients between 2004 and 2013, 45% of prescriptions for antibiotics were made without any clear rationale. Twenty-eight percent of antibiotics were prescribed without evidence of seeing the provider, and 17% were given without documentation for infection-related diagnosis. Inappropriate antibiotic prescribing leads to antibiotic resistance on an individual as well as community levels, particularly against public health threats such as carbapenem-resistant Enterobacteriaceae and methicillin-resistant *Staphylococcus aureus*.^14^ Furthermore, overuse of antibiotics increases the risk of adverse effects such as rash, GI upset, and renal dysfunction.^6^ The risk of infection with *Clostridioides difficile* increases with duration of exposure to antibiotics.^15-16^ A recent systematic review of 160 observational studies focused on antibiotic exposure and long-term health outcomes in childhood showed that postnatal antibiotic exposure was associated with asthma (odd ratio = 2.0), wheezing (OR= 1.8), allergic rhinoconjunctivitis (OR= 1.7), food allergies (OR=1.4), being overweight (OR= 1.2) and obesity (OR = 1.2).^17^

State level differences in antibiotic prescribing are well known.^18-19^ Geographic areas with high antibiotic consumption were associated with increased antibiotic resistance, and that broad-spectrum antibiotics were more likely to be associated with antibiotic resistance. The South census region had the highest prescribing rate in the US.^18-19^ Kentucky had the highest prescriptions rates (1,281 prescriptions per 1,000 persons) which was about four-fold higher than Alaska (348 per 1,000 persons).^18^ An analysis of nationally representative data from the Medical Expenditure Panel Surveys (MEPS) revealed relatively stable antibiotic use overall from 2000 (382/1,000 persons) to 2010 (384/1,000) but a doubling for broad spectrum antibiotics.^20^ Relative to 27 European countries, the US ranked fourth in 2004 for daily doses of antibacterials and was number one for azithromycin, levofloxacin, and cefdinir.^21^

Due to the public health risks that arise from antibiotic resistance,^1,6^ and other adverse health outcomes associated with unnecessary exposure to antibiotics,^17^ it is imperative to be continuously vigilant to identify and avoid potentially unnecessary prescriptions. This study examined the temporal profile of outpatient antibiotic use as reported by MEPS and geographical patterns of antibiotic prescribing rates among US Medicaid program beneficiaries.

## METHODS

### Participants

The MEPS is a national set of surveys of patients, families, and their medical providers with a sample of 35,000 respondents to estimate health care use annually which has been employed in prior research.^20^Medicaid is a joint federal and state program that provides coverage for 75 million people or 21% of the US population.^22^ It is one of the largest payers for health care in the U.S. All states provide coverage for outpatient prescription drugs.^22^

### Procedures

Antibiotic agents that were consistently ranked in the top three-hundred most prescribed were examined from 2013-2022. These antibiotics included amoxicillin, amoxicillin-clavulanate, azithromycin, cephalexin, ciprofloxacin, doxycycline, levofloxacin, and trimethoprim-sulfamethoxazole were examined from 2013-2020 (the most recent available when data-analysis was completed in December, 2022).

Quarterly state antibiotic prescription totals and population data during 2018-2019 were collected from the Medicaid State Drug Utilization database. These years were selected because they were relatively recent but also before any disruptions associated with the COVID-19 pandemic. Antibiotics were identified using the generic and trade names.^23^ Antibiotics were further categorized as broad or narrow spectrum based on their potential for influencing antibacterial resistance as well as their spectrum of activity according to the NCQA *Antibiotics of Concern* list. Azithromycin, levofloxacin, moxifloxacin, and ciprofloxacin were categorized as broad-spectrum antibiotics, while amoxicillin, cephalexin, doxycycline, and trimethoprim sulfamethoxazole were categorized as narrow-spectrum.^24,25^ These agents were selected based on overlap with those listed in MEPS and past Medicaid investigations.^18^

The states were divided into geographic regions according to the US Census (Supplementary Figure 1).

### Data Analysis

The prescriptions as well as patients in MEPS were extracted. The overall total was calculated for each year and for broad and narrow-spectrum agents. After the total number of prescriptions was collected and prescription rate per 1,000 Medicaid enrollees was calculated, we input the data into IMB SPSS. Pearson correlations were completed between antibiotics for each year. Although r > .279 was significant (*p* < .05) for associations with fifty states, correlations above .50 were considered large. We created figures and heatmaps through Statistical Analysis System‘s John‘s Macintosh Project (SAS JMP) and GraphPad Prism to show variation across regions in the United States. Variability was reported as the standard error of the mean (SEM). A *p*-value of < .05 was considered statistically significant.

## RESULTS

### MEPS

During the study period, there were 687,979,538 million antibiotic prescriptions dispensed in the US. Total antibiotics decreased by −38.7% while the US population increased by 5.2% from 2013 to 2020. Broad spectrum agents had a larger decline (−53.7%) than narrow spectrum (−23.5%). More specifically, amoxicillin decreased by −39.1%, trimethorprim-sulfamethoxazole by −25.9% and cephalexin by −8.9%, while there was in increase in doxycycline (+16.2%). Azithromycin (−68.3%), levofloxacin (−56.4%), and ciproflaxin (−33.8%) experienced pronounced declines while amoxicillin-clavulante increased (+3.9%, Figure 1A).

**Figure 1.**
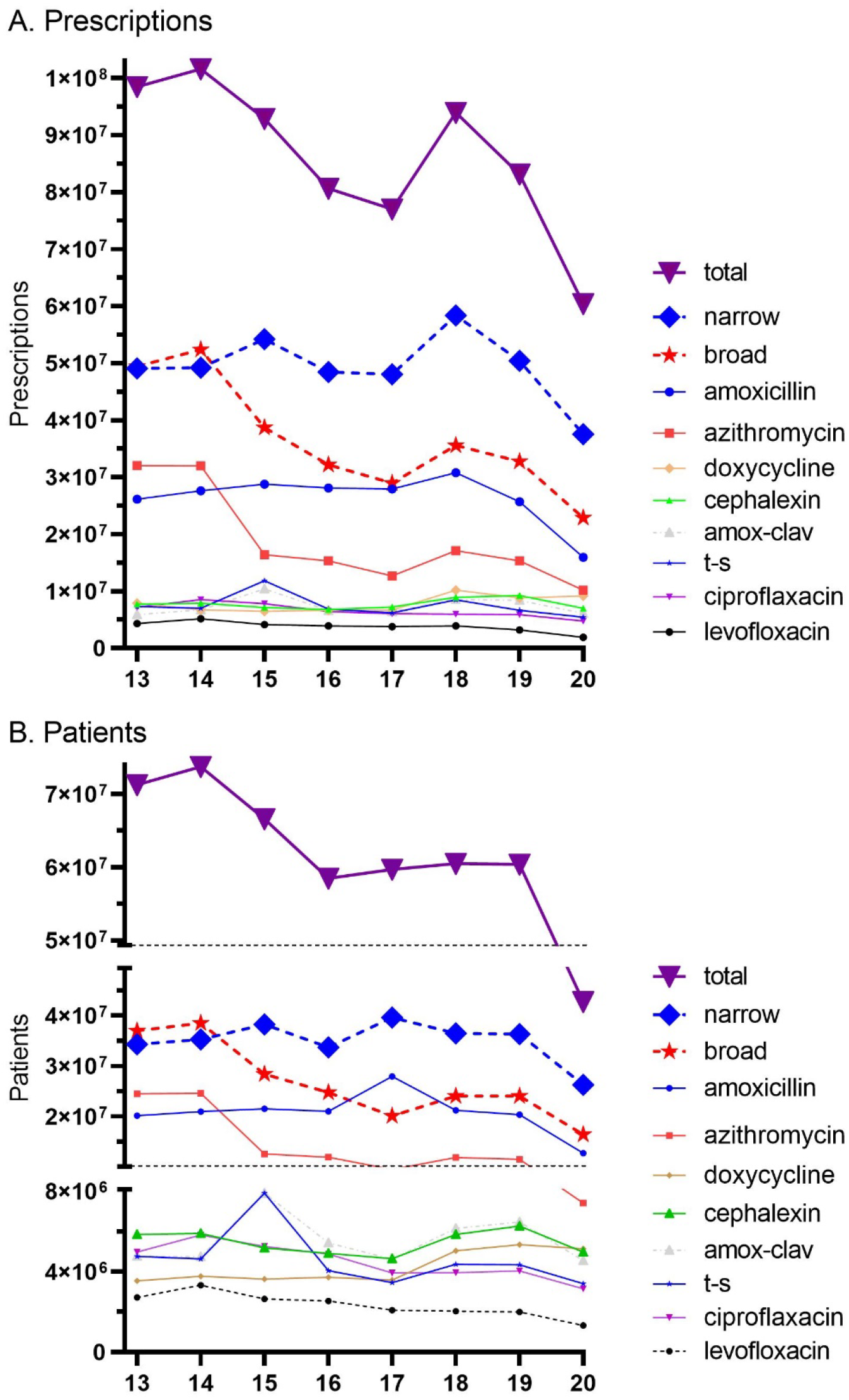
Total prescriptions (A) and patients (B) prescribed narrow and broad spectrum antibiotics as reported by the Medical Expenditure Panel Survey.

Similarly, the total number of patients that were dispensed antibiotics decreased from 71.3 million in 2013 to 42.7 million (−40.0%) in 2020. The reduction in recipients of broad-spectrum agents (−55.6%) was over twice as large as narrow spectrum (−23.3%).

### Medicaid

The total amount of prescriptions in 2019 (33,011,946 prescriptions) was 9.61% lower than in 2018 (36,519,951 prescriptions). Similarly, prescriptions per 1,000 enrollees in 2019 (464) were 6.47% lower than in 2018 (494). Broad spectrum antibiotics decreased by 14.11% relative to 7.95% for narrow spectrum agents. Figure 1 shows the quarterly prescriptions. Amoxicillin was the most prescribed antibiotic, accounting for almost half (47.0% of the total), followed by azithromycin (18.76%), cephalexin (11.80%), trimethoprim sulfamethoxazole (9.25%), doxycycline (6.42%), ciprofloxacin (4.39%), levofloxacin (1.93%), and moxifloxacin (0.49%). Azithromycin and amoxicillin showed the most dynamic quarterly changes with highest levels in the first and fourth quarters (Figure 2).

**Figure 2.**
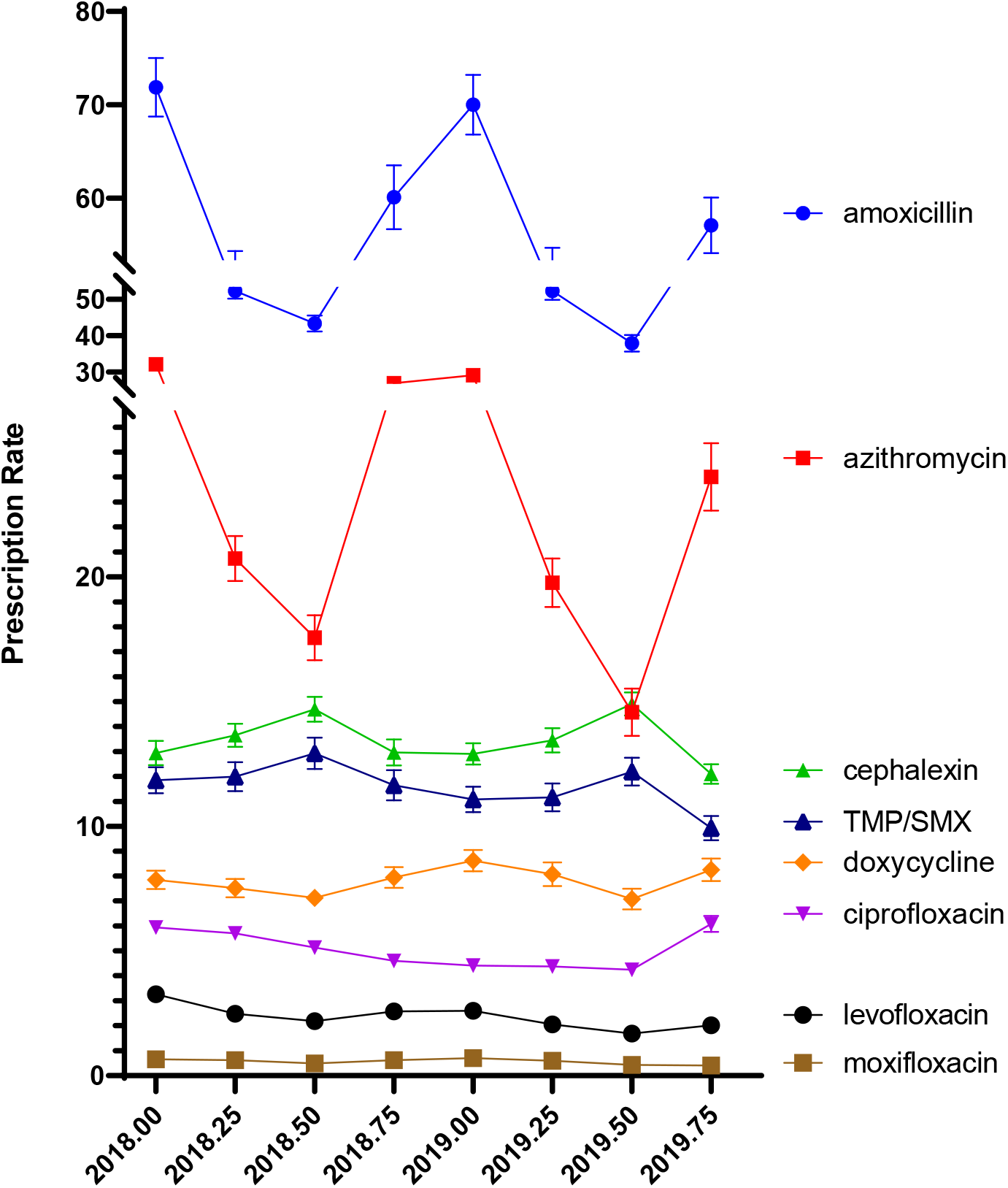
Quarterly antibiotic prescribing per 1,000 Medicaid patients.

**Figure 3.**
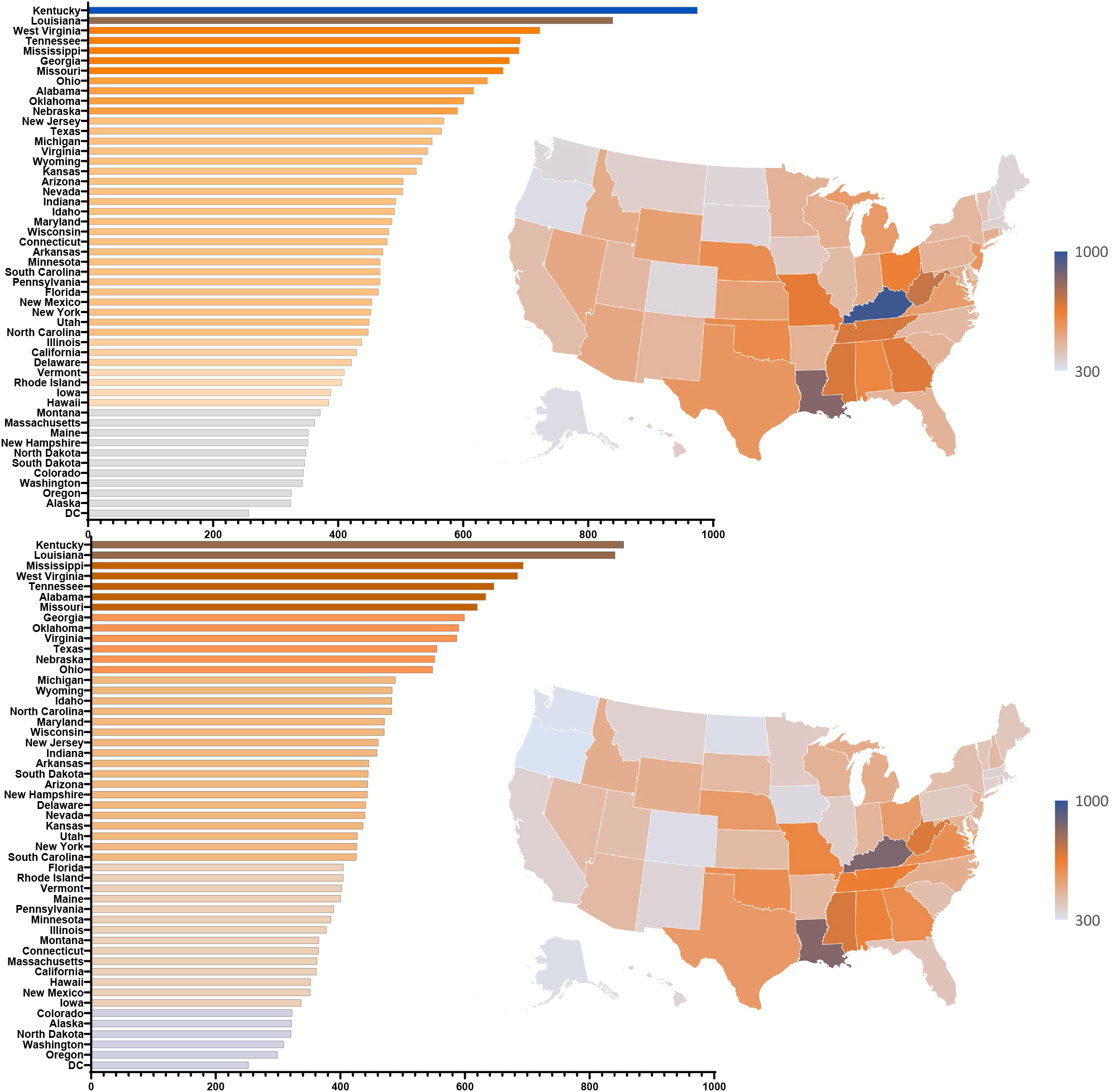
Total antibiotic prescribing rate per state in 2018 (top) and 2019 (bottom) per 1,000 Medicaid patients.

**Figure 4.**
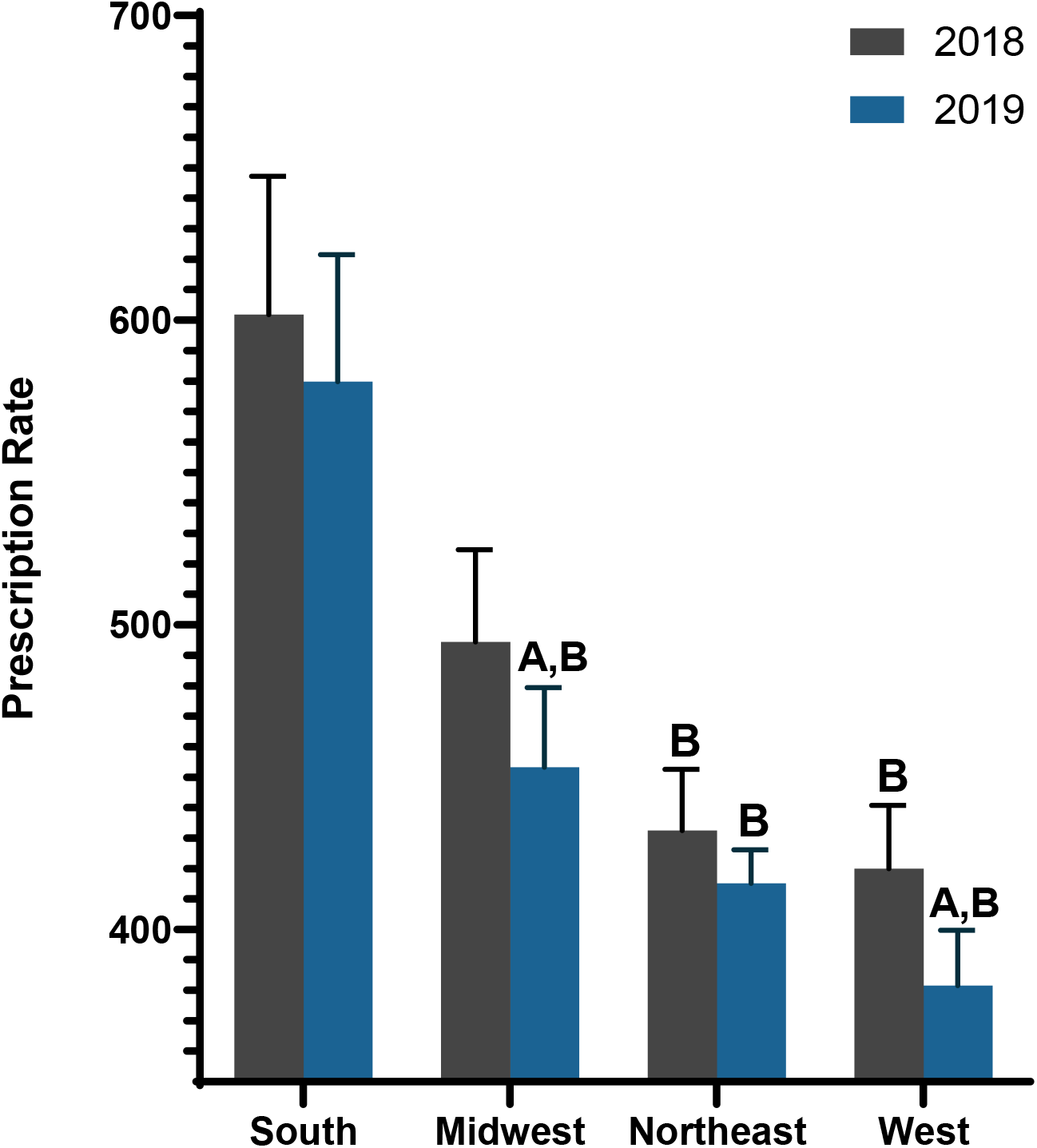
Antibiotic prescribing per 1,000 Medicaid enrollees by census regions. A indicates statistically significant difference within the region. B indicates statistically significant difference compared to the South.

There was a 3-fold difference between the highest (Kentucky = 975/1,000) and lowest prescribing states (Alaska = 325/1,000) in 2018 (Figure 2A). Similarly, there was a 2.86-fold difference between the highest (Kentucky = 855/1,000) and lowest (Oregon = 299/1,000) states in 2019 (Figure 2B). Nationally, there was some evidence for a West-South gradient with 7 of the 13 western states making up the ten lowest prescribing states: Oregon (50th), Washington (49^th^), Alaska (47th), Colorado (46^th^), New Mexico (44^th^), Hawaii (43^rd^), California (42^nd^), and 9 of the 15 southern states making up the ten highest prescribing states: Kentucky (1^st^), Louisiana (2^nd^), Mississippi (3^rd^), Tennessee (4^th^), Alabama (6^th^) Georgia (8^th^), Oklahoma (9^th^), Virginia (10^th^) in 2019. Similarly, Alaska (50^th^), Oregon (49th), Washington (48th), and Colorado (47^th^) were relatively low prescribing states in 2018.

Further examination of geographical patterns was made by grouping states by census regions. The South prescribed 28.80% more antibiotics (590/1,000) than the West (420/1,000) in 2018. Similarly, the South prescribed 35.57% more antibiotics (563/1,000) than the West (363/1,000) in 2019. <expand> From 2018 to 2019, the South decreased by −4.51% which was smaller than the declines in the Midwest (−11.21%), Northeast (−9.05%), and West (−13.58%)

The correlation between population corrected antibiotic prescriptions in 2018 is shown in Table1. The correlation between the total was significantly associated with most individual agents except for moxifloxacin and this pattern was replicated in 2019. There were strong correlations between levofloxacin and other antibiotics, again, with the exception of moxifloxacin, for both 2018 and 2019. This same pattern of strong associations was also identified for trimethoprim-sulfamethoxazole.

## DISCUSSION

There are at least three key findings from this pharmacoepidemiological report of US MEPS and Medicaid patients. First, there were reductions in overall dispensing nationally. A national Medicaid reduction of −6.47% in antibiotic prescribing was observed with −7.95% for narrow spectrum and −14.11% for broad spectrum agents from 2018 to 2019. These changes were generally less pronounced than the reductions for azithromycin (−10.5%), doxycycline (−13.6%), amoxicillin (−16.5%), levofloxacin (−17.7%) and trimethoprim-sulfamethoxazole (−21.7%) from 2018 to 2019 in the MEPS. Looking more broadly in MEPS (i.e. 2013 to 2020), outpatient prescribing decreased −38.7% with broad spectrum (−53.7%) agents declining more than narrow spectrum (−23.5%). These results are quite different than an earlier MEPS report which determined that broad spectrum antibiotics doubled with a 2.5-fold increase for those aged 18-49 from 2000 to 2010.^20^ There are many differences between Medicaid and commercially insured patients. A study from Massachusetts found that Medicaid insured parents were more trusting of the information about coughs, colds and the flu from product advertisements, social media, and other parents but less trusting than commercially insured parents of the Centers for Disease Control, the Department of Public Health, and their child‘s doctor.^25^

Second, there were sizable regional and pronounced state-level variation in antibiotic prescribing. This included a 2.8-fold difference between the highest (Kentucky and Louisiana) and lowest (Alaska and Oregon) states. These findings are consistent and extend upon past geographical findings.^7,26^ Analysis of QuintilesIMS reports from 2013 determined that children aged ≤ 19 from Louisiana and Mississippi received four-fold more azithromycin prescriptions than children from Oregon or Alaska.^26^ Similarly, examination of IQVIA database revealed 2.7 fold more antibiotic prescriptions in 2019 in Mississippi (1,193/1,000 patients) than Alaska (447/1,000 patients).^7^ Kentucky children from the more rural and eastern Knott County received three-fold more antibiotic prescriptions than more urban children from Louisville.^27^ Elevations in southern relative to western states are not limited to Medicaid insured patients.^18,28,29^ However, Medicare Part D patients in the South in 2013 had 1,623 antibiotics per 1,000 patients) which was only 26% more claims than those in the West (1,292).^29^

Third, and perhaps most novel, was the finding that there were several strong (r > .50) associations between state-level use of different antibiotics in 2018 which were generally very consistent when examined for the subsequent year. The state in which a patient resides in (e.g. West Virginia versus adjacent Virginia) might be anticipated to have only slight biological or bacteriological impact but could be quite important due to the many social determinants of health. Counties with higher incomes and education prescribed fewer antibiotics while those with more obese adults, health care providers, and females prescribed more antibiotics in 2011.^18^

Also of note was that antibiotic prescribing varied based on the quarter in the year. The highest antibiotic prescribing rates occurred found during October to March, while prescribing rates fell from April to the end of September. This is consistent with patterns in upper respiratory tract infections which are often more prevalent during fall or winter months and decrease during warmer months.

There are some strengths and limitations to these two complementary databases. First, the existence of state level disparities among Medicaid programs is only suggestive of inappropriate antibiotic prescribing which has been identified previously.^10-13^ Further research is necessary to determine if these state-level disparities persist, or were exacerbated, during the COVID-19 pandemic. Second, only eight antibiotics were examined for MEPS and eight for Medicaid. However, prior investigations have noted that the top five antibiotics accounted for 87% of all prescriptions.^28^ The MEPS and Medicaid databases are based on claims and do not account for antibiotics that were diverted from other sources.

## Conclusions & Relevance

This study provides an overview of antibacterial prescribing practices for US outpatients and in the Medicaid system. Key findings include disparities in antibacterial prescribing rates among different regions across the US including southeastern states. In conjunction with prior investigations which have characterized the risk factors for heightened prescribing, we are cautiously optimistic that these findings will contribute to renewed antibiotic stewardship efforts.

## Data Availability

All data produced are available online at https://data.medicaid.gov/

## Acknowledgements

Thanks to Sean Kane, PharmD for making MEPS information available through ClinCalc.com and Kenneth McCall, PharmD for feedback on an earlier version of this manuscript.

---

**Table 1.**
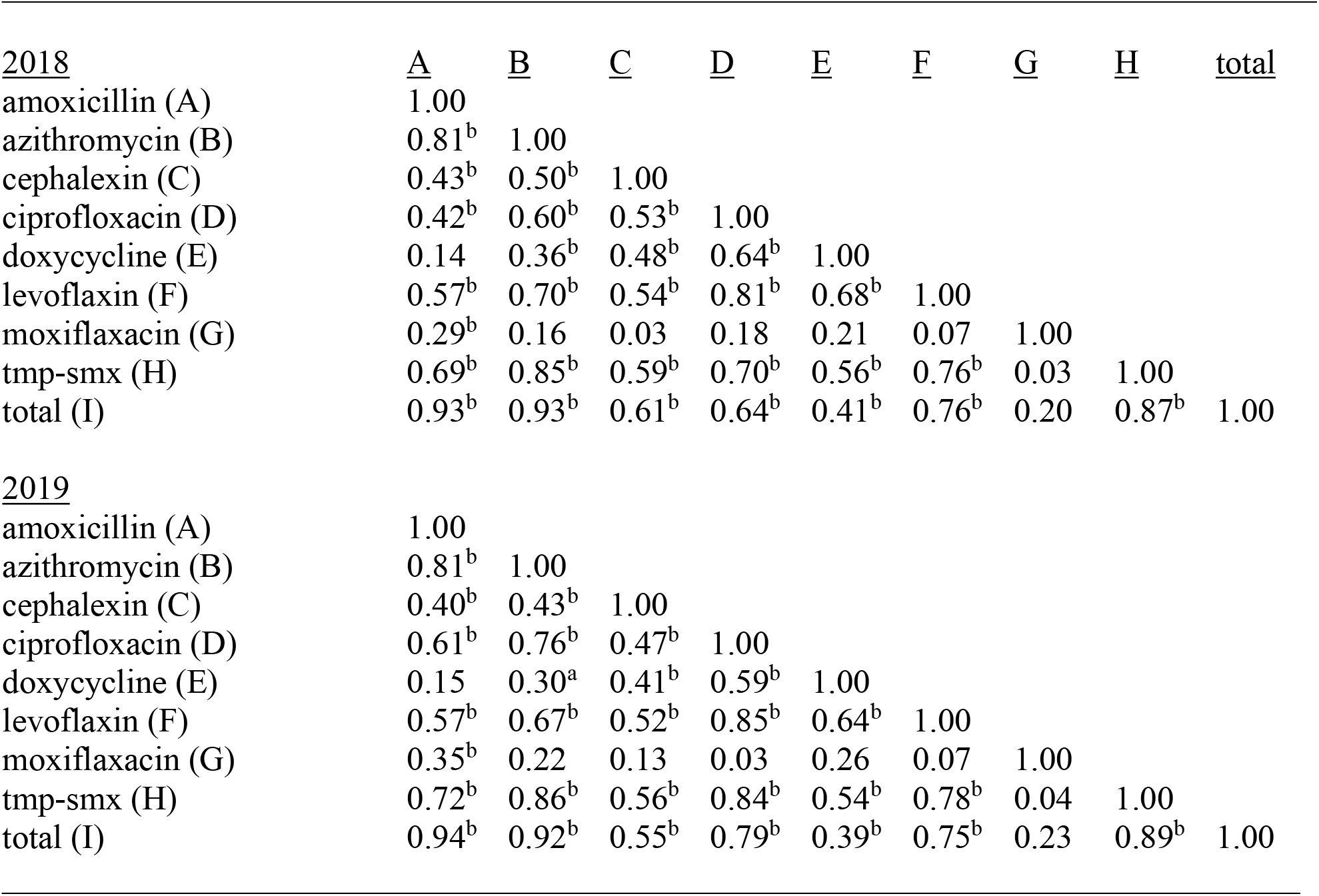
Correlation matrix showing associations between antibiotic prescribing to Medicaid patients in 2018 (top) and 2019 (bottom). trimethoprim sulfamethoxazole: tmp-smx. ap < .05 (.279), bp<.01(.361)

| <u>2018</u> | <u>A</u> | <u>B</u> | <u>C</u> | <u>D</u> | <u>E</u> | <u>F</u> | <u>G</u> | <u>H</u> | <u>total</u> |
| --- | --- | --- | --- | --- | --- | --- | --- | --- | --- |
| amoxicillin (A) | 1.00 |  |  |  |  |  |  |  |  |
| azithromycin (B) | 0.81<sup>b</sup> | 1.00 |  |  |  |  |  |  |  |
| cephalexin (C) | 0.43<sup>b</sup> | 0.50<sup>b</sup> | 1.00 |  |  |  |  |  |  |
| ciprofloxacin (D) | 0.42<sup>b</sup> | 0.60<sup>b</sup> | 0.53<sup>b</sup> | 1.00 |  |  |  |  |  |
| doxycycline (E) | 0.14 | 0.36<sup>b</sup> | 0.48<sup>b</sup> | 0.64<sup>b</sup> | 1.00 |  |  |  |  |
| levofloxacin (F) | 0.57<sup>b</sup> | 0.70<sup>b</sup> | 0.54<sup>b</sup> | 0.81<sup>b</sup> | 0.68<sup>b</sup> | 1.00 |  |  |  |
| moxifloxacin (G) | 0.29<sup>b</sup> | 0.16 | 0.03 | 0.18 | 0.21 | 0.07 | 1.00 |  |  |
| tmp-smx (H) | 0.69<sup>b</sup> | 0.85<sup>b</sup> | 0.59<sup>b</sup> | 0.70<sup>b</sup> | 0.56<sup>b</sup> | 0.76<sup>b</sup> | 0.03 | 1.00 |  |
| total (I) | 0.93<sup>b</sup> | 0.93<sup>b</sup> | 0.61<sup>b</sup> | 0.64<sup>b</sup> | 0.41<sup>b</sup> | 0.76<sup>b</sup> | 0.20 | 0.87<sup>b</sup> | 1.00 |
| <u>2019</u> |  |  |  |  |  |  |  |  |  |
| amoxicillin (A) | 1.00 |  |  |  |  |  |  |  |  |
| azithromycin (B) | 0.81<sup>b</sup> | 1.00 |  |  |  |  |  |  |  |
| cephalexin (C) | 0.40<sup>b</sup> | 0.43<sup>b</sup> | 1.00 |  |  |  |  |  |  |
| ciprofloxacin (D) | 0.61<sup>b</sup> | 0.76<sup>b</sup> | 0.47<sup>b</sup> | 1.00 |  |  |  |  |  |
| doxycycline (E) | 0.15 | 0.30<sup>a</sup> | 0.41<sup>b</sup> | 0.59<sup>b</sup> | 1.00 |  |  |  |  |
| levofloxacin (F) | 0.57<sup>b</sup> | 0.67<sup>b</sup> | 0.52<sup>b</sup> | 0.85<sup>b</sup> | 0.64<sup>b</sup> | 1.00 |  |  |  |
| moxifloxacin (G) | 0.35<sup>b</sup> | 0.22 | 0.13 | 0.03 | 0.26 | 0.07 | 1.00 |  |  |
| tmp-smx (H) | 0.72<sup>b</sup> | 0.86<sup>b</sup> | 0.56<sup>b</sup> | 0.84<sup>b</sup> | 0.54<sup>b</sup> | 0.78<sup>b</sup> | 0.04 | 1.00 |  |
| total (I) | 0.94<sup>b</sup> | 0.92<sup>b</sup> | 0.55<sup>b</sup> | 0.79<sup>b</sup> | 0.39<sup>b</sup> | 0.75<sup>b</sup> | 0.23 | 0.89<sup>b</sup> | 1.00 |

**Supplementary Figure 1.**
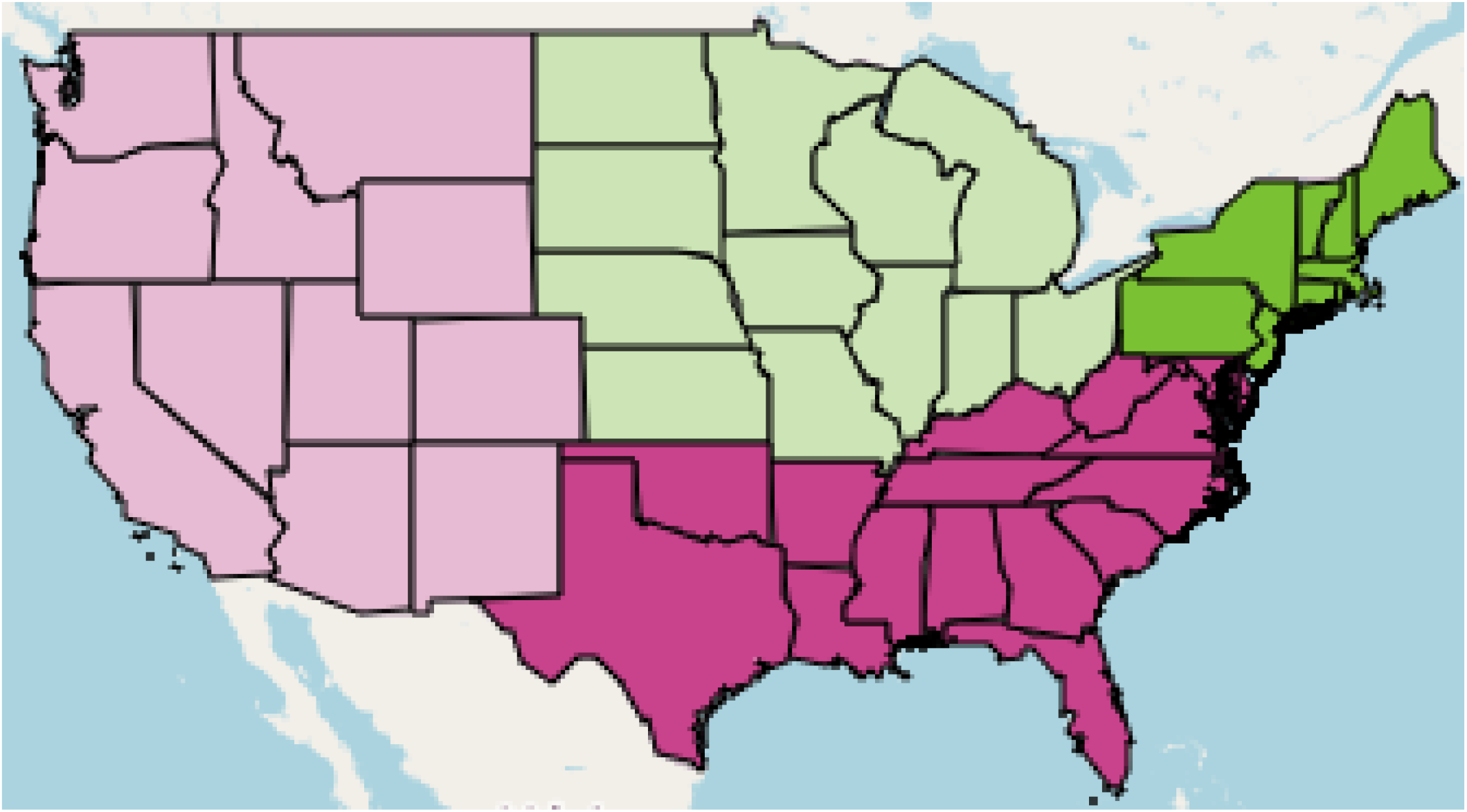
US Census regions including the **West, Midwest, South**, and **Northeast**. Alaska and Hawaii are not shown. Image created with http://www.heatmapper.ca/geomap/

